# Neuroanatomical Deficits in Visual Cortex Subregions of Individuals with Psychosis Spectrum Disorders linked to Symptoms, Cognition, and Childhood Trauma

**DOI:** 10.1101/2025.02.12.25322031

**Authors:** Halide Bilge Türközer, Victor Zeng, Dung Hoang, Jothini Sritharan, Neha Iska, Elena I. Ivleva, Brett A. Clementz, Godfrey D. Pearlson, Sarah Keedy, Elliot S. Gershon, Carol A. Tamminga, Matcheri S. Keshavan, Paulo Lizano

**Author notes:** Address correspondence to Halide Bilge Türközer, M.D., at and/or Paulo Lizano, M.D., Ph.D., at. denotes shared senior authorship (Matcheri S. Keshavan, Paulo Lizano).

## Abstract

**Objective:** The visual system is a significant site of pathology in psychosis spectrum disorders. However, there is limited research investigating human visual cortex (VC) subregions in this population. Using data from the Bipolar-Schizophrenia Network on Intermediate Phenotypes Consortium (BSNIP-1, BSNIP-2, PARDIP), this study examined structural measures in VC subregions in individuals with psychosis spectrum disorders.

**Methods:** Cortical surface area and thickness in five VC subregions (hOc1, hOc2, hOc3v, hOc4v, MT) were quantified using FreeSurfer v7.1.0 and compared between individuals with psychosis (*n*=1211) and healthy controls (*n*=734). Regional specificity was examined by controlling for total surface area or mean cortical thickness. ComBat was used to harmonize scanner effects. Associations between VC measures and symptom severity, cognition, and childhood trauma scores were assessed.

**Results:** Individuals with psychosis demonstrated smaller surface area in hOc1, hOc2, and hOc3v, and lower cortical thickness in all five VC subregions compared to healthy controls. Thickness reductions in hOc1, hOc4v, and MT were regionally specific. hOc4v and MT were among the top three regions exhibiting the most robust cortical thickness deficits (*d* = −0.38 to −0.40) across all VC and Desikan-Killiany brain regions. Lower thickness in mid-level visual subregions were associated with greater positive symptoms, poorer cognition, and higher childhood trauma scores.

**Conclusions:** This study demonstrates that the visual cortex is among the most profoundly affected brain regions in psychotic disorders. Different patterns of area and thickness changes across early and mid-level visual subregions, along with their varying associations with clinical measures, suggest distinct developmental and disease-related influences.

## INTRODUCTION

The visual system is a significant site of pathology in psychosis spectrum disorders. Initially, basic visual symptoms and visual perceptual impairments were the sole focus of attention (1–3). However, recent studies have demonstrated structural, functional, and molecular changes in the visual cortex (VC) (4–7), along with retinal changes (8). These findings indicate that the range of visual system impairments in psychotic disorders is much wider than previously recognized. However, the extent of visual system impairments, their clinical significance, and the reproducibility of these findings are yet to be determined and present a critical gap in the field.

One question that remains underexplored is the extent of VC changes in psychosis spectrum disorders. Early studies with small sample sizes demonstrated smaller cortical thickness and lower surface area in the occipital cortex (9, 10). Research on VC subregions is even more scarce (10–12). This is an important question because VC subregions have distinct structure, function, and developmental trajectories (13). Furthermore, VC subregions are differentially impacted by some risk factors associated with psychotic disorders, such as childhood trauma (14). Therefore, examining the changes in VC subregions may provide information about the mechanisms and developmental processes associated with psychotic disorders.

Of the limited studies that investigated VC subregions in psychosis spectrum disorders, one study showed no changes in V1, V2, or V3 structure (10), while another demonstrated smaller cortical thickness in V2 and V5/middle temporal (MT)+, but not in V1 (12). The largest study to date (4), published by our group, demonstrated area, thickness, and volume reductions in V1, V2, and MT in individuals with psychosis spectrum disorders, along with area and volume reductions in MT in their first-degree relatives using the Bipolar-Schizophrenia Network on Intermediate Phenotypes (B-SNIP-1) dataset. Area and volume reductions in V1 and V2 were sex-dependent, affecting only females with psychosis. Furthermore, structural changes in VC subregions were associated with poor general cognition, worse response inhibition, and increased C-reactive protein levels.

Building on our previous work, this study aimed to investigate VC subregions in greater detail using a larger dataset and a new cytoarchitectonic atlas of the human ventral visual stream. Using harmonized BSNIP-1, BSNIP-2, and Psychosis and Affective Research Domains and Intermediate Phenotypes (PARDIP) datasets, in individuals with psychosis spectrum disorders (IwP) and healthy controls (HC), we examined gray matter surface area and cortical thickness in five VC subregions. These comprised human occipital cytoarchitectonic area 1 (hOc1), hOc2, ventral hOc3 (hOc3v), ventral hOc4 (hOc4v), and middle temporal area (MT) using FreeSurfer’s cytoarchitectonic vcAtlas (15) and Broadman Area maps (16). Our secondary aim was to evaluate the associations between VC measures and symptom severity, cognitive measures, and childhood trauma scores (only in BSNIP-2 and PARDIP participants). We hypothesized that compared to HC, IwP demonstrate lower surface area and cortical thickness in VC subregions, with more pronounced surface area changes in early VC regions and more prominent cortical thinning in mid-level VC regions. We expected associations between lower mid-level VC structural measures and greater symptoms, worse cognition, and higher childhood trauma.

## METHODS AND MATERIALS

### Study Sample and Clinical Assessments

Participants with available structural brain imaging data from BSNIP-1, BSNIP-2, and PARDIP datasets were included in this study. B-SNIP and PARDIP protocols, detailed elsewhere (17, 18), were approved by local Institutional Review Boards across study sites. All participants provided informed consent. The Structured Clinical Interview for DSM-IV Axis I disorders (SCID-IV) was used to assess for current and lifetime psychiatric diagnoses (19). 1211 individuals with psychosis spectrum disorders (Schizophrenia, n=469; Schizoaffective Disorder, n=373; or Psychotic Bipolar Disorder, n=369) and 734 healthy participants were included in the final analysis (BSNIP-1, N=798; BSNIP-2, N=1028; PARDIP, N=119; **Table S1**). Psychosis symptom severity was assessed using the Positive and Negative Syndrome Scale (PANSS) (20). Brief Assessment of Cognition in Schizophrenia (BACS) was administered to assess cognition (21). Childhood Trauma Questionnaire (CTQ) was used to assess childhood trauma in BSNIP-2 and PARDIP (22). See **Table 1** for demographic and clinical characteristics.

**Table 1:** Demographic and Clinical Measures.

|  | HC (n=734) |  | lwP (n=1211) |  | p-value |
| --- | --- | --- | --- | --- | --- |
|  | N | % | N | % |  |
| Sex (Female) | 425 | 57.9 | 605 | 50 | <0.001 |
| Race |  |  |  |  | 0.010 |
| African American | 227 | 31 | 454 | 37 |  |
| Caucasian | 402 | 55 | 612 | 51 |  |
| Other | 105 | 14 | 145 | 12 |  |
| Diagnosis |  |  |  |  |  |
| Schizophrenia |  |  | 469 | 39 |  |
| Schizoaffective Disorder |  |  | 373 | 31 |  |
| Bipolar Disorder with Psychotic Features |  |  | 369 | 30 |  |
|  | Mean | SD | Mean | SD |  |
| Age | 36.05 | 12.3 | 37.1 | 12.1 | 0.065 |
| GAF | 84.8 | 6.8 | 54.1 | 13.1 | <0.001 |
| SFS total | 154.6 | 16.9 | 124.2 | 24.3 | <0.001 |
| Daily CPZ equivalent | - |  | 534.3 | 748.2 | - |
| Age of onset | - |  | 19.4 | 8.4 |  |
| PANSS total | - |  | 61.9 | 19.1 | - |
| PANSS Positive total | - |  | 15.9 | 6.1 | - |
| PANSS Negative total | - |  | 14.9 | 6.2 | - |
| PANSS General total | - |  | 31.2 | 9.5 | - |
| CTQ Total | 44.98 | 10.03 | 59.77 | 18.64 | <0.001 |
| CTQ Emotional Abuse | 7.46 | 3.62 | 12.12 | 5.78 | <0.001 |
| CTQ Physical Abuse | 6.74 | 2.67 | 9.4 | 4.81 | <0.001 |
| CTQ Sexual Abuse | 5.92 | 2.88 | 9.57 | 6.67 | <0.001 |
| CTQ Emotional Neglect | 8.61 | 4.12 | 12.33 | 5.35 | <0.001 |
| CTQ Physical Neglect | 6.31 | 2.12 | 8.78 | 3.91 | <0.001 |

**Note:** HC = healthy control; lwP = Individuals with Psychotic Spectrum Disorders; SD = standard deviation; CPZ = chlorpromazine; GAF = Global Assessment of Functioning; SFS = Birchwood Social Functioning Scale; PANSS = Positive and Negative Syndrome Scale; CTQ = Childhood Trauma Questionnaire.

### Structural Magnetic Resonance Imaging

T1-weighted 3T images were acquired at seven sites (Baltimore, MD; Boston, MA; Chicago, IL; Dallas, TX; Athens, GA; Hartford, CT; Detroit, MI) based on the Alzheimer’s Disease Neuroimaging Initiative protocol (http://adni.loni.usc.edu). FreeSurfer version 7.1.0. was used for processing. Scans were quality checked and manually edited to improve the segmentation of cortical regions by a trained rater (VZ). The Desikan-Killiany Atlas segmentation (23) was also inspected for motion artifacts and for inaccurate gray/white matter segmentation, which were manually corrected. Gray matter measures for hOc1 (similar to BA17/V1), hOc2 (similar to BA18/V2), hOc3v (follows hOc2 ventrally and is located in the collateral sulcus), and hOc4v (located anterior to hOc3v, on the ventral aspect of the occipital lobe) were obtained using the vcAtlas map, which is a cross-validated cytoarchitectonic atlas of the human ventral visual stream which is available in FreeSurfer 7.1.0 (15). Gray matter measures for MT were obtained using FreeSurfer’s Broadmann Area atlas, which was derived from postmortem brains and surface-based analysis to localize the Broadman Areas (16). Visual cortical segmentation maps were also visually inspected for correct segmentation.

Since different scanners were used at the different sites, there were significant scanner-dependent differences for thickness and surface area measures in all regions. To harmonize across scanners, ComBat from the Surrogate Variable Analysis package in R (https://rdrr.io/bioc/sva/man/ComBat.html) was applied to VC measures, DK Atlas measures and total intracranial volume. Outliers were examined and all data points which deviated more than 4 standard deviations within a site were removed (N=12 for VC regions, N=81 for DK Atlas regions). Diagnostic group, sex, and age were used as covariates to preserve biological differences in the data and to account for imbalances in sample demographics. The measures were normalized before ComBat adjustment. Almost all significant scanner differences were eliminated for the various measures (see **Supplement**).

### Statistical Analysis

Statistical analyses were performed in SPSS version 28.0 (IBM Corp.) and R version 4.1.3. Demographic variables were compared across groups using chi square tests or student t-tests. For group comparisons (IwP vs. HC), analyses of covariance were performed for mean area and thickness measures in hOc1, hOc2, hOc3v, hOc4v, and MT using ComBat-adjusted total intracranial volume (for area) as a covariate (model A). To assess regional specificity − whether the structural changes observed were specific to certain regions rather than reflecting global brain alterations − group comparisons were repeated using total surface area or mean cortical thickness as covariates (model B). Age, sex, and race were included as covariates in both models A and B. With separate ANCOVAs, we explored group-by-sex interactions for these measures, using age, race, and total intracranial volume (TIV) (only for surface area) as covariates.

To compare the magnitude of visual cortical changes to other brain regions, surface area and cortical thickness differences were explored in 58 DK Atlas brain regions in IwP vs. HC using analyses of covariance. Age, sex, race, and intracranial volume (for surface area) were used as covariates. Bilateral DK Atlas occipital cortex regions (lingual gyrus, pericalcarine, cuneus, and lateral occipital cortices) and middle temporal gyrus were excluded from this analysis given that they overlap with VC regions explored in this study.

Effect sizes were calculated using Cohen’s f2 and Cohen’s d. The false discovery rate (FDR) approach was used to correct for multiple testing (*q* value), was applied separately for area and thickness, and statistical significance was set at *q* = 0.05. For VC regions, FDR corrections included group comparisons from five subregions (mean hOc1, hOc2, hOc3v, hOc4v, and MT). For all brain regions, corrections included groups comparisons from 29 bilateral DK Atlas regions and five bilateral VC regions (68 total regions). We used one-way ANOVA’s to compare the effect size (Cohen’s *d*) of each significant area and thickness difference to the effect sizes of changes seen in other brain regions.

Relationships between VC measures and current symptoms, cognition, and childhood trauma scores were assessed using partial Spearman correlations given that several clinical and cognitive measures were non-normally distributed. Age, race, sex, and total intracranial volume (for area) were included as covariates. FDR corrections were applied separately for clinical (five VC subregions X four PANSS measures), cognitive measures (five VC subregions X seven standardized BACS scores), and childhood trauma scores (five VC subregions X six CTQ scores).

## RESULTS

### Structural changes in VC subregions in IwP compared to HC (Model A)

IwP demonstrated lower mean surface area in hOc1 (*d* = −0.15), hOc2 (*d* = −0.12), and hOc3v (*d* = −0.1), (**Figure 1A, Table S2**), and smaller cortical thickness in all five VC subregions (hOc1, *d* = −0.1; hOc2, *d* = −0.2; hOc3v, *d* = −0.26; hOc4v, *d* = −0.38; MT, *d* = −0.44) compared to HCs (**Figure 1B, Table S3**). In IwP, there were no VC measure differences across the three diagnostic groups (schizophrenia, schizoaffective disorder, and psychotic bipolar disorder) or BSNIP Biotypes (Supplement). There were no FDR significant group-by-sex interactions.

**Figure 1.**
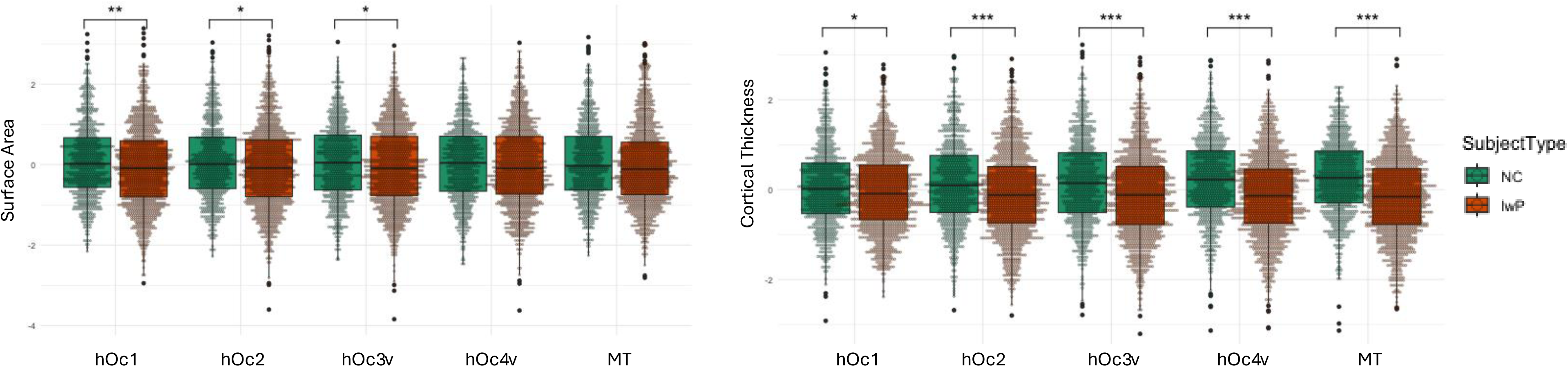
Comparing visual cortical area and thickness across groups. Boxplot with individual ComBat-adjusted data points demonstrating group comparisons for **(A)** surface area (hOc1, hOc2, hOc3v, hOc4v, MT), **(B)** cortical thickness (hOc1, hOc2, hOc3v, hOc4v, MT) across healthy controls (HC) and individuals with psychosis spectrum disorders (IwP). Panel (A) shows significantly lower area in hOc1 (*q* = 0.005, *d* = − 0.15), hOc2 (q = 0.020, *d* = − 0.12), and hOc3v (q = 0.040, *d* = − 0.10) in IwP compared with HC subjects. Panel (B) shows significant cortical deficits in hOc1 (*q* = 0.031, *d* = − 0.10), hOc2 (*q* < 0.001, *d* = − 0.20), hOc3v (*q* < 0.001, *d* = − 0.26), hOc4v (*q* < 0.001, *d* = − 0.38), and MT (*q* < 0.001, *d* = − 0.44) in IwP compared to HC. \**q* < 0.05, **q < 0.01, ***q < 0.001.

### Regional specificity of structural VC changes (Model B)

There were no regional surface area changes. However, lower cortical thickness in hOc1 (*q* = 0.03, *d* = −0.11), hOc4 (*q* = 0.001, *d* = −0.16), and MT (*q* < 0.001, *d* = −0.20) were regionally specific (Table S4). There were no significant group-by-sex interactions.

### Correlations with clinical measures

In IwP, lower MT thickness was associated with higher PANSS Positive scores (*r* = −0.10, *q* = 0.02) and PANSS Total scores (trend-level, *r* = −0.08, *q* = 0.06). There were no associations between VC measures and antipsychotic dosage (daily chlorpromazine equivalents).

In all participants (IwP and HC together), lower hOc4v and MT thickness values were associated with higher CTQ Total scores, along with Emotional, Physical, and Sexual Abuse subscores (**Table 2**). Additionally, lower MT thickness was correlated with higher Emotional and Physical Neglect subscores (**Table 2**). When investigated separately in IwP and HC, no correlations between VC measures and childhood trauma scores survived FDR correction.

**Table 2:** Correlations between Clinical/Cognitive Measures and VC Structural Measures.

|  |  |  |  |  |  |
| --- | --- | --- | --- | --- | --- |
| **Clinical/Cognitive Measure** | **Group** | **VC Measure** | **r-value** | **p-value** | **q-value** |
| PANSS Positive | lwP | MT Thickness | - 0.101 | 0.001 | 0.020 |
| CTQ Total | All participants | hOc4v Thickness | - 0.10 | 0.001 | 0.004 |
| CTQ Total | All participants | MT Thickness | - 0.143 | <0.001 | <0.001 |
| CTQ Emotional Abuse | All participants | hOc4v Thickness | - 0.075 | 0.012 | 0.036 |
| CTQ Emotional Abuse | All participants | MT Thickness | - 0.116 | <0.001 | <0.001 |
| CTQ Physical Abuse | All participants | hOc4v Thickness | - 0.078 | 0.009 | 0.030 |
| CTQ Physical Abuse | All participants | MT Thickness | - 0.126 | <0.001 | <0.001 |
| CTQ Sexual Abuse | All participants | hOc4v Thickness | - 0.100 | 0.001 | 0.004 |
| CTQ Sexual Abuse | All participants | MT Thickness | - 0.125 | <0.001 | <0.001 |
| CTQ Emotional Neglect | All participants | MT Thickness | - 0.102 | 0.001 | 0.004 |
| CTQ Physical Neglect | All participants | MT Thickness | - 0.089 | 0.003 | 0.011 |
| BACS Total | All participants | hOc3v Thickness | 0.053 | 0.023 | 0.049 |
| BACS Total | All participants | hOc4v Thickness | 0.107 | <0.001 | <0.001 |
| BACS Total | All participants | MT Thickness | 0.155 | <0.001 | <0.001 |
| BACS Total | All participants | MT Area | 0.079 | 0.001 | 0.018 |
| BACS Total | lwP | MT Thickness | 0.094 | 0.001 | 0.018 |
| Verbal Memory | All participants | hOc4v Thickness | 0.075 | 0.001 | 0.004 |
| Verbal Memory | All participants | MT Thickness | 0.092 | <0.001 | <0.001 |
| Digit Sequencing | All participants | hOc3v Thickness | 0.058 | 0.013 | 0.030 |
| Digit Sequencing | All participants | hOc4v Thickness | 0.062 | 0.008 | 0.020 |
| Digit Sequencing | All participants | MT Thickness | 0.115 | <0.001 | <0.001 |
| Digit Sequencing | All participants | MT Area | 0.080 | 0.001 | 0.018 |
| Token Motor | All participants | hOc4v Thickness | 0.087 | <0.001 | <0.001 |
| Token Motor | All participants | MT Thickness | 0.104 | <0.001 | <0.001 |
| Token Motor | All participants | MT Area | 0.065 | 0.005 | 0.040 |
| Verbal Fluency | All participants | hOc4v Thickness | 0.074 | 0.001 | 0.004 |
| Verbal Fluency | All participants | MT Thickness | 0.087 | <0.001 | <0.001 |
| Symbol Coding | All participants | hOc4v Thickness | 0.102 | <0.001 | <0.001 |
| Symbol Coding | All participants | MT Thickness | 0.156 | <0.001 | <0.001 |
| Tower of London | All participants | hOc4v Thickness | 0.086 | <0.001 | <0.001 |
| Tower of London | All participants | MT Thickness | 0.121 | <0.001 | <0.001 |
| Tower of London | All participants | MT Area | 0.068 | 0.004 | 0.044 |
| Tower of London | lwP | MT Thickness | 0.102 | 0.001 | 0.018 |

**Note:** lwP = Individuals with Psychotic Spectrum Disorders; PANSS = Positive and Negative Syndrome Scale; CTQ = Childhood Trauma Questionnaire; BACS = Brief Assessment of Cognition in Schizophrenia. Partial Spearman correlations were performed between clinical measures (PANSS Total score and subscores, CTQ Total score and subscores, BACS Total score and subscores) and hOc1, hOc2, hOc3v, hOc4v, and MT surface area and cortical thickness measures. Correlations were performed using age, sex, race, and total intracranial volume (only for area) as covariates. q-value=FDR-corrected p value. Only measures surviving the multiple comparison correction are reported in the table.

### Correlations with cognitive measures

In all participants, lower hOc4v and MT thickness values were correlated with poorer BACS Total score and all six subscores (**Table 2**). Additionally, lower hOc3v thickness was associated with poorer BACS Total scores and BACS Digit Sequencing subscore. Smaller MT surface area was correlated with poorer BACS Total scores, along with BACS Digit Sequencing, Token Motor, and Tower of London subscores (**Table 2**).

In IwP, lower MT thickness was correlated with poorer BACS Total scores (*r* = 0.09, *q* = 0.018) and BACS Tower of London subscores (*r* = 0.1, *q* = 0.018) **(Table 2)**. There were no correlations between BACS scores and VC measures in HC.

### Effect sizes of changes in VC measures in comparison to other brain regions

When group comparisons for 68 brain regions were examined, bilateral hOc1 and right hOc2 were among the eleven brain regions (eight distinct regions) that showed significant surface area differences between IwP and HC (Supplement). The effect sizes in these three VC regions (*d* = −0.14 to −0.15) were comparable to those observed in other brain regions with significant surface area deficits (*d* = −0.12 to −0.17) (one-sample T tests, p<0.05) (**Figure 2**). Left hOc1 (*d* = −0.15) was among the top three regions with the most robust surface area deficits.

**Figure 2.**
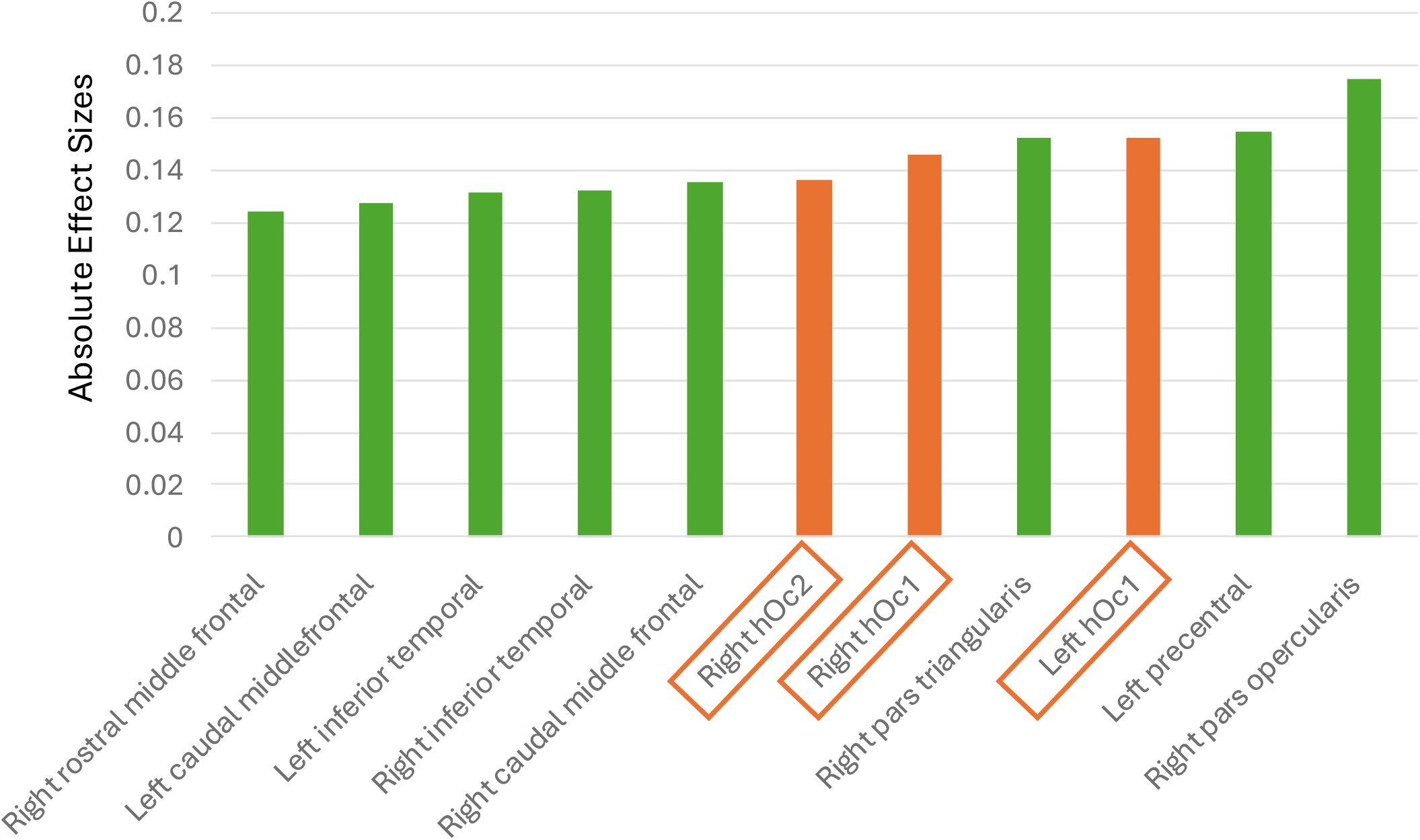
Effect sizes (all negative, but displayed in absolute values) in visual cortex (VC) subregions and Desikan-Killiany (DK) brain regions that show significant surface area deficits (*d* = −0.12 to −0.17) in individuals with psychosis spectrum disorders compared to healthy controls, following FDR correction. A total of 68 regions were examined, including 29 bilateral DK Atlas regions and five bilateral VC subregions, excluding five DK regions that overlap with VC subregions.

Most DK Atlas and all VC regions, except for right hOc1, demonstrated significant thickness differences between IwP and HC (*q* <0.05). Bilateral MT and right hOc4 (*d* = −0.38 to −0.40), along with bilateral fusiform regions (*d*’s = −0.40), were the top five regions (top three distinct regions) showing the most robust cortical thickness deficits across the 68 brain regions analyzed (*d* = −0.11 to −0.40), with significantly larger effect sizes than the other regions (one-sample t tests, p < 0.001) (**Figure 3**).

**Figure 3.**
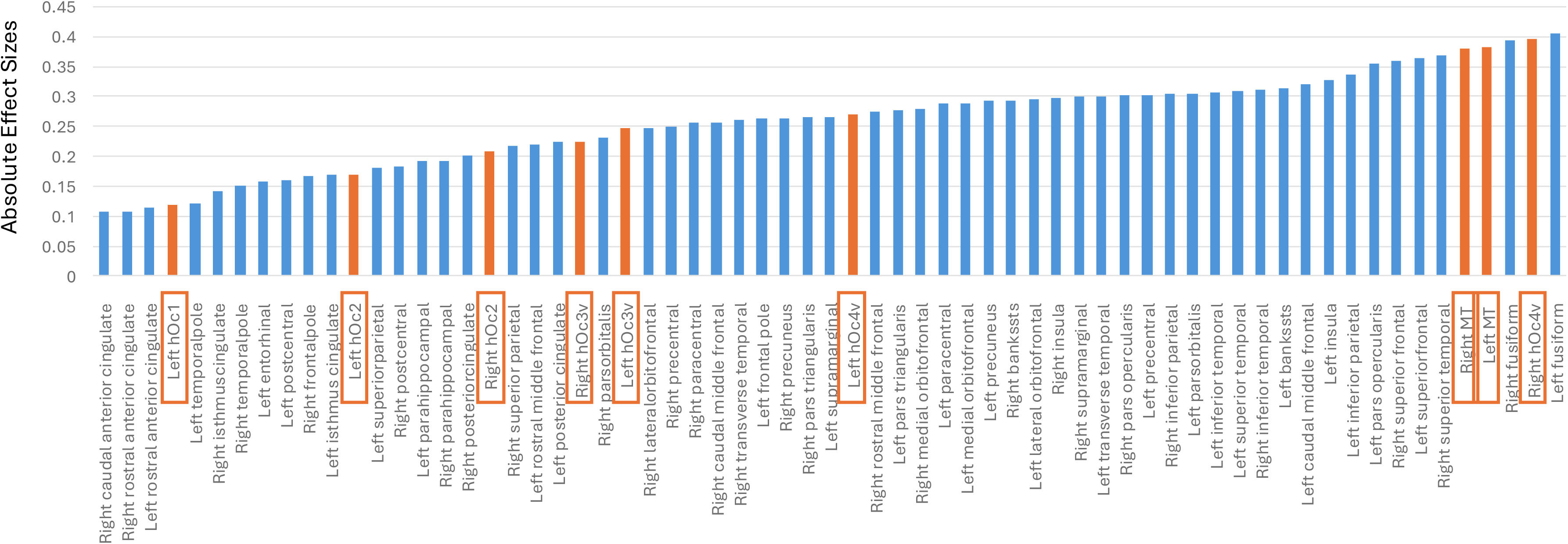
Effect sizes (all negative, but displayed in absolute values) in visual cortex (VC) subregions and Desikan-Killiany (DK) brain regions that show significant cortical thickness deficits (*d* = −0.11 to −0.40) in individuals with psychosis spectrum disorders compared to healthy controls, following FDR correction. A total of 68 regions were examined, including 29 bilateral DK Atlas regions and five bilateral VC subregions, excluding five DK regions that overlap with VC subregions.

## DISCUSSION

This study demonstrates surface area deficits in early-level VC subregions (hOc1, hOc2, hOc3v) and cortical thickness deficits in all VC subregions (hOc1, hOc2, hOc3v, hOc4v, and MT) in individuals with psychosis spectrum disorders, with more robust and regional thickness deficits in the mid-level VC subregions (hOc4v, MT). Notably, hOc1 and hOc2 were among the only eight regions that showed reduced surface area across the entire brain. hOc4v and MT were among the top three regions with the most robust cortical thickness deficits across all brain regions. Among VC subregions, structural changes primarily in mid-level VC subregions (hOc4v, MT) were associated with clinical and cognitive measures. These results showcase the significance of visual cortical deficits in psychotic disorders, standing out against the widespread cortical changes seen across the brain. Distinct patterns of area and thickness changes seen in VC subregions, along with their varying associations with clinical and cognitive symptoms, indicate the influence of distinct developmental and disease-related processes driving this differentiation.

This study offers a more detailed examination of visual cortical alterations in psychotic disorders within a larger dataset, addressing gaps in the limited literature on this topic. Our group’s previous findings from the BSNIP-1 dataset showed area and thickness deficits in BA17/V1, BA18/V2, and MT in IwP (n=530) compared to HC (n=323) (4). Surface area deficits were more prominent in V1 and V2, with sex-specific effects, and cortical thinning was more robust in MT. In this larger harmonized dataset of BSNIP-1, BSNIP-2, and PARDIP studies, the more granular data from five VC subregions allowed for a finer investigation of these VC alterations. This study demonstrates that surface area changes are predominantly confined to early VC subregions (hOc1, hOc2, hOc3v), with the most pronounced alteration in hOc1. In contrast, there is a progressive worsening in cortical thickness from hOc1 (*d* = −0.1) to MT (*d* = −0.44), with MT having the strongest associations with clinical and cognitive measures.

This marked contrast between surface area and thickness patterns in VC subregions aligns with the evidence that these measures offer distinct and complementary information (24–26). Surface area and thickness are driven by distinct cellular mechanisms, have distinct genetic etiologies and developmental trajectories (24, 27). Evidence suggests that surface area is influenced by genes that are critical to early brain development, while cortical thickness is influenced by genes that govern later developmental processes, such as synaptic pruning and myelination (24, 28, 29). Cortical thickness is also considered more plastic in later stages of life, influenced by environmental, disease-related, and neurodegenerative factors (28, 30, 31). Evidence from psychosis research aligns with these findings. Large-scale studies, including an ENIGMA meta-analysis, showed that cortical thinning in schizophrenia is associated with the disease and its severity, while surface area changes were not associated with clinical symptoms (9, 30). On the other hand, surface area appears to be more strongly affected by familial and genetic factors in psychotic disorders (32, 33). This is in line with our previous findings from the BSNIP-1 dataset, which also included first degree relatives of individuals with psychosis (4). We demonstrated that V1 and V2 surface area measures had the highest familiality among all VC measures. Taken together, our results suggest that visual cortex regions (hOc1, hOc2) that develop earlier are affected by early developmental factors related to psychosis (e.g. familial/hereditary factors), thus showing greater surface area deficits, while later-developing higher-level visual regions (hOc4v, MT) are affected by disease-related processes and potentially environmental factors that are active later in life, resulting in more robust thickness deficits.

In the context of global cortical alterations seen in psychotic disorders, one critical question is whether these structural VC changes hold particular significance. Here, we not only demonstrated the regional specificity of thickness changes in VC subregions, but also showed that hOc4v and MT were among the top three regions exhibiting the most robust cortical thickness deficits across the entire brain, following the fusiform gyrus. This is in line with findings from the ENIGMA meta-analysis on cortical structural changes in schizophrenia (30). Middle temporal gyrus (quantified using the DK Atlas) was among the regions that show the greatest thickness reduction in this meta-analysis. Furthermore, our results demonstrated that hOc1 and hOc2 were among the only eight regions that showed surface area changes in the entire brain, with hOc1 being among the top three regions with the most robust surface area deficits. These results differ from the findings of the ENIGMA meta-analysis, which demonstrated widespread surface area deficits in the brain (30). This difference could be explained by the ENIGMA metaanalysis’ larger sample size or methodological differences between the two study. Please see the Supplement for a more detailed discussion of this point. Overall, our results highlight that the visual cortex is among the most profoundly affected brain regions in psychotic disorders.

Our findings demonstrate an association between thinner hOc4v and MT and poorer cognition in IwP. MT, a region of the visual association cortex and the dorsal (“where”) pathway, plays a significant role in spatial processing, motion perception, integration, and segmentation (34). It is one of the regions in the dorsal attention network (35), which is involved in verbal and visual memory tasks (36). hOc4v (corresponds to ventral human V4) is involved in color selectivity, along with the encoding of texture, form, surfaces (37), and attentional modulation of spatial tuning (38). Our results are consistent with previous studies showing impairments in dorsal pathway functions (e.g. motion perception, spatial attention, eye movements) in psychotic disorders and at-risk states, and their association with higher-level cognitive deficits (2, 39). Together, this evidence indicates a connection between impairments in mid-level visual regions and the higher-level cognitive deficits observed in psychotic disorders. This relationship warrants further investigation, especially considering its potential implications for developing novel treatments targeting cognitive symptoms in psychotic disorders. Could mid-level visual regions serve as potential mechanistic targets for cognitive treatments? There are ongoing studies that employ visual remediation and interventional treatments to investigate this critical question, with promising preliminary results (40–43).

MT thickness was also associated with psychosis symptom severity, particularly with positive symptoms. This finding aligns with recent studies on networks involved in visual hallucinations (VH) using lesion network mapping methods. These studies showed that bilateral extrastriate visual cortices (beVC), which includes bilateral MT regions, were causally linked to visual hallucinations (44–46). Our group expanded on these findings by conducting a pilot study targeting the beVC (including MT) using high-definition transcranial direct current stimulation (HD-tDCS) in stable outpatients with psychosis spectrum disorders (47, 48). We showed that HD-tDCS over the beVC reduced P1 amplitude (generated in beVC) and PANSS General scores. The change in P1 amplitude was related to the change in symptoms, demonstrating target engagement that could predict clinical outcomes (47). In the same individuals, treatment with HD-tACS (alternating current) over the beVC (MT) resulted in improvements in digit sequencing, verbal fluency, and tower of London tasks (41). This targeting approach was expanded to a 71-year-old woman with treatment resistant and distressing VH. In this case study, HD-tDCS resulted in a rapid, consistent and sustained reduction in symptoms (49). These brain stimulation findings demonstrate the translational neuroscience potential of the current study.

The association between thinner hOc4v and MT and childhood trauma is also noteworthy and aligns with previous studies. The visual cortex is increasingly recognized as a primary region impacted by childhood trauma (14, 50, 51). Cortical thickness and volume deficits have been reported in primary visual and visual association cortices in young individuals with exposure to childhood trauma (50, 51). Among individuals who witnessed domestic violence during childhood, Tomoda et al. also demonstrated lower thickness in MT in those who presented with psychiatric symptoms (e.g. depression, anxiety, trauma-related symptoms) compared to relatively resilient subjects with no mental health symptoms (51). Along with exposure to past trauma, growing evidence suggests that acute trauma exposure and resultant post-traumatic symptoms are associated with structural changes in the ventral visual stream (52, 53), which includes the hOc4v region. Our findings contribute to the growing body of evidence indicating the visual system’s involvement in the brain’s response to trauma and point to a potential shared site of pathology in the development of psychosis and trauma-related disorders.

There were a few notable limitations to the current study. One limitation is that the psychosis group was largely composed of chronically ill, medicated individuals. Even though there were no associations between anti-psychotic use and VC measures, potential confounders related to chronic medication effects and medical comorbidities cannot be eliminated. Additionally, although we considered several major confounders (age, sex, race, intracranial volume), there are other potential confounding factors (e.g. lifetime antipsychotic exposure, substance abuse, comorbid medical illness, body mass index, eye-related metrics) that were not measured in this study.

In conclusion, we demonstrated distinct patterns of surface area and cortical thickness changes across early and mid-level visual cortex subregions in individuals with psychotic disorders. In this large multicenter sample, we also showed that the visual cortex is among the most profoundly affected brain regions in individuals with psychotic disorders. These findings not only reinforce the evidence that the visual system is significantly affected in psychotic disorders but also highlight it as one of the primary sites of pathology. Future research investigating the mechanistic relationships between visual system alterations and psychotic symptoms may inform the development of novel predictive tools, such as risk and prognostic markers, as well as targeted therapeutic interventions for psychotic disorders.

## Data Availability

All data produced in the present study are available upon reasonable request to the authors

## Acknowledgments

This study was funded by National Institute of Health, National Institute of Mental Health (NIMH): MH096942, MH078113, MH096900, MH103366, MH096913, MH077851, MH096957, MH077945, MH077852, MH077862, MH103368. HBT was supported by Harvard Medical School’s Dupont Warren and Livingston Fellowships, McLean Hospital’s Pope-Hintz Fellowship, and the Brain and Behavior Research Foundation’s NARSAD Young Investigator Grant. SK was supported by the University of Chicago Magnetic Resonance Imaging Research Center (S10OD018448). PL was supported by the NIMH (K23MH122701).

The authors express gratitude to the patients who contributed their time and effort to participate in this study. Additionally, they thank the numerous researchers and clinicians who assisted in recruitment and data collection.

