## Supplement for "Neuroanatomical Deficits in Visual Cortex Subregions of Individuals with Psychosis Spectrum Disorders linked to Symptoms, Cognition, and Childhood Trauma"

**1. ComBat analysis**

Surface area and thickness measures across the VC and DK regions showed significant scanner differences before ComBat adjustment. ComBat was applied separately to area and thickness measures. After applying ComBat, all scanner-dependent differences got eliminated in the VC regions. Almost all scanner-dependent differences got eliminated in the DK regions, except for the following measures: right lateral orbitofrontal area, left frontal thickness, right frontal thickness, left superior frontal thickness, right superior frontal thickness, left mean thickness, right mean thickness, and mean thickness.

**2. Comparisons of VC measures across BSNIP Biotypes in individuals with psychotic disorders (IwP)**

The lack of differences in visual cortex measures across DSM diagnoses (schizophrenia, schizoaffective disorder, and bipolar 1 disorder with psychotic features) is consistent with previous BSNIP studies showing overlapping biomarker profiles across these three disorders (Tamminga et al., *Am J of Psychiatry*, 2013; Ivleva et al., *Am J Psychiatry*, 2013). Previously, BSNIP consortium developed three biomarker-driven disease constructs, Biotypes 1-3. These Biotypes represent biologically distinct subgroups of individuals with psychosis based on cognitive and neurophysiological biomarkers, independent of clinical phenomenology (Clementz et al., *Am J Psychiatry*, 2016). Given the lack of differences across DSM diagnoses, we explored the differences across Biotypes in IwP. There were no significant differences in hOc1, hOc2, hOc3v, hOc4v, or MT measures across the three Biotypes.

**3. Effect sizes of cortical changes in all brain regions**

To compare the magnitude of change in VC measures to other brain regions, we assessed cortical differences in 68 brain regions between IwP and HC, using ANCOVA. Bilateral DK Atlas occipital cortex regions (lingual gyrus, pericalcarine, cuneus, and lateral occipital cortices) and middle temporal gyrus were excluded from this analysis given that they overlap with VC regions explored in this study. FDR correction was applied for group comparisons from 68 regions (29 bilateral DK Atlas regions and five bilateral VC regions). IwP demonstrated significant surface area reductions in 11 regions (*d* = −0.12 to −0.17, mean *d* = −0.142, SD=0.015) and cortical thinning in 65 regions (*d* = −0.11 to −0.40, mean *d* = −0.259, SD=0.077). The largest effect sizes were observed for right pars opercularis surface area (*d* = −0.17, q=0.007) and left fusiform thickness (*d* = −0.405, q<0.001).

Our results demonstrating that hOc1 and hOc2 were among the only eleven regions (eight distinct regions) that showed surface area changes in the entire brain differ from the findings of the ENIGMA meta-analysis, which demonstrated widespread surface area deficits in the brain (31). This could be due to our inclusion of total intracranial volume (TIV) as a covariant in our analysis, while Van Erp et al. did not make this adjustment. FreeSurfer recommends correcting for TIV in surface area analyses to account for inter-individual differences in head size (https://surfer.nmr.mgh.harvard.edu/fswiki/eTIV). To explore this potential explanation, we repeated the group comparison analyses for surface area measurements of DK Atlas regions without adjusting for TIV. This analysis showed 51 DK Atlas regions with significant surface area changes (*d* = −0.10 to −0.21), similar to the findings of ENIGMA. Of note, there were no significant TIV differences between individuals with psychosis and healthy controls in our sample. These findings suggest that, when accounted for head size using TIV, surface area changes in individuals with psychotic disorders are limited, with visual cortex being among the few affected regions. Overall, our results highlight that the visual cortex is among the most profoundly affected brain regions in psychotic disorders.

| **Table S1: Sample sizes by Diagnoses and Study** | | | | |
| --- | --- | --- | --- | --- |
| **Group** | **BSNIP-1** | **BSNIP-2** | **PARDIP** | **Total** |
| Schizophrenia | 207 | 262 | 0 | 469 |
| Schizoaffective Disorder | 125 | 248 | 0 | 373 |
| Psychotic Bipolar Disorder | 177 | 139 | 53 | 369 |
| Healthy Controls | 289 | 379 | 66 | 734 |
| Total | 798 | 1028 | 119 | 1945 |

| **Table S2: ANCOVA statistics for hOc1, hOc2, hOc3v, hOc4v, and MT surface area measures in IwP vs. HC** | | | | |
| --- | --- | --- | --- | --- |
| **Region** | **F-value** | **q-value** | **Cohen’s *f*^2^** | **Cohen’s d** |
| Mean hOc1 area | *F*(1,1938)=11.608 | **0.005** | 0.006 | -0.15 |
| Mean hOc2 area | *F*(1,1938)=7.150 | **0.020** | 0.004 | -0.12 |
| Mean hOc3v area | *F*(1,1938)=5.067 | **0.040** | 0.003 | -0.10 |
| Mean hOc4v area | *F*(1,1938)=0.550 | 0.459 | 0.0002 | -0.03 |
| Mean MT area | *F*(1,1938)=2.351 | 0.156 | 0.001 | -0.07 |
| **Note:** Total intracranial volume, sex, race, and age were included as covariates. q= FDR corrected p values. Bold values denote statistical significance at the q<0.05 level. | | | | |

| **Table S3: ANCOVA statistics for hOc1, hOc2, hOc3v, hOc4v, and MT cortical thickness measures in IwP vs. HC** | | | | |
| --- | --- | --- | --- | --- |
| **Region** | **F-value** | **q-value** | **Cohen’s *f*^2^** | **Cohen’s d** |
| Mean hOc1 thickness | *F*(1,1939)=4.644 | **0.031** | 0.002 | -0.10 |
| Mean hOc2 thickness | *F*(1,1939)=20.040 | **<0.001** | 0.01 | -0.20 |
| Mean hOc3v thickness | *F*(1,1939)=33.910 | **<0.001** | 0.017 | -0.26 |
| Mean hOc4v thickness | *F*(1,1939)=71.264 | **<0.001** | 0.037 | -0.38 |
| Mean MT thickness | *F*(1,1939)=94.511 | **<0.001** | 0.049 | -0.44 |
| **Note:** Sex, race, and age were included as covariates. q= FDR corrected p values. Bold values denote statistical significance at the q<0.05 level. | | | | |

| **Table S4: ANCOVA statistics for hOc1, hOc2, hOc3v, hOc4v, and MT cortical thickness measures in IwP vs. HC (Regional Specificity Analysis – Model B)** | | | | |
| --- | --- | --- | --- | --- |
| **Region** | **F-value** | **q-value** | **Cohen’s *f*^2^** | **Cohen’s d** |
| Mean hOc1 thickness | *F*(1,1938)=5.384 | **0.034** | 0.003 | -0.10 |
| Mean hOc2 thickness | *F*(1,1938)=0.681 | 0.409 | 0.0004 | -0.04 |
| Mean hOc3v thickness | *F*(1,1938)=0.743 | 0.409 | 0.0004 | -0.04 |
| Mean hOc4v thickness | *F*(1,1938)=11.975 | **0.001** | 0.006 | -0.16 |
| Mean MT thickness | *F*(1,1938)=94.511 | **<0.001** | 0.01 | -0.20 |
| **Note:** Sex, race, age, and mean cortical thickness were included as covariates. q= FDR corrected p values. Bold values denote statistical significance at the q<0.05 level. | | | | |
